# Anatomic and Functional Outcomes of Lamellar Macular Hole Surgery: Predictive Factors and Associated Complications

**DOI:** 10.1101/2025.01.09.25320019

**Authors:** Yosra Er-reguyeg, Sihame Doukkali, Mélanie Hébert, Eunice Linh You, Serge Bourgault, Mathieu Caissie, Éric Tourville, Ali Dirani

**Affiliations:** Faculty of Medicine, Laval University, Quebec City, Quebec, Canada; Department of Ophthalmology, Hospital Saint-Sacrement, Laval University, Quebec City, Quebec, Canada

## Abstract

**Purpose:** To analyze the anatomic and functional outcomes of lamellar macular hole (LMH) surgery.

**Patients and methods:** This is a retrospective interventional cohort study of ninety patients with unilateral idiopathic LMH who underwent pars plana vitrectomy (PPV) with membrane peeling for LMH between 2014 and 2021. We evaluated the anatomic and functional success of PPV with membrane peeling for treating LMH, compared surgical outcomes between the two LMH subtypes (“true” LMH and epiretinal foveoschisis (ERMF)), and identified predictive factors for anatomical and functional success. Primary outcomes included final postoperative best-corrected visual acuity (BCVA) and LMH closure. Variables associated with final BCVA were assessed using a multiple linear regression model.

**Results:** 51 subjects presented with ERMF, while 39 presented with “true” LMH. LMH closure occurred in 80 cases. “True” LMH cases had a lower rate of closure (“true” LMH closure rate: 76.9%, vs. ERMF closure rate: 94.2%, p=0.005) and were more at risk of developing a postoperative macular hole (p=0.008). A significant difference was observed between median [Q1, Q3] preoperative BCVA (0.42 [0.26, 0.61]) and final BCVA (0.31 [0.14, 0.48], p=0.024). “True” LMH without epiretinal proliferation (β=0.194, p=0.040) was associated with worse final BCVA in multivariate analysis.

**Conclusion:** Results support the effectiveness of PPV as a treatment for LMH. “True” LMHs had worse anatomic outcomes than ERMFs.

## Introduction

Lamellar macular hole (LMH) is characterized as a partial-thickness defect in the inner layers of the fovea. Since J.D. Gass first documented the disease in 1975, the diagnostic criteria for LMH changed considerably.^1^ Established as the gold standard in LMH assessment, the emergence of spectral domain optical coherence tomography (SD-OCT) led to new diagnostic criteria including an irregular foveal contour, a break in the inner fovea, dehiscence of the inner foveal retina from the outer retina and absence of a full-thickness foveal defect with preservation of foveal photoreceptors.^2^

Govetto et al. later divided LMHs into “tractional” and “degenerative” subtypes.^3^ Recently, Hubschman et al. separated lesions previously called LMH into two entities: epiretinal membrane foveoschisis (ERMF) and “true” LMH.^4^ ERMF, which is comparable to what was previously considered as tractional LMH, occurs in the presence of a contractile epiretinal membrane (ERM) and a foveoschisis at the level of the Henle’s fiber layer (HFL), two mandatory diagnosis criteria. Three optional criteria were also suggested: the presence of microcystoid spaces in the inner nuclear layer (INL), retinal thickening, and retinal wrinkling.

In contrast to ERMF, “true” LMH, which is comparable to what was previously considered degenerative LMH, requires the presence of an irregular foveal contour, a foveal cavity with undermined edges, and at least one other sign evoking loss of foveal tissue. Associated pathological changes can include epiretinal proliferation (ERP), foveal bump, and ellipsoid zone (EZ) disruption.

It is unclear whether ERMF and “true” LMH are two distinct entities, as the latest findings suggest that both originate from a tractional event and that the ERP often found in “true” LMH may be a repair process derived from a tractional impairment to the foveolar Müller cells.^5–9^ Moreover, Su et al.’s findings indicate that in LMHs displaying ERP, epiretinal traction is commonly observed.^10^

Disagreement remains regarding the surgical treatment of LMH involving integral membrane peeling. While surgery may prevent visual acuity (VA) loss and further deterioration of the foveal profile, some studies report outcomes that vary based on the morphological features of the LMH.^11,12^ The fovea-sparing and flap embedding techniques are new methods recently developed to address complications commonly associated with standard integral membrane peeling, showing promising postoperative outcomes. However, standard peeling remains widely used in LMH surgery.^13,14^ Thus, we aimed to assess the anatomic and functional outcomes of LMH surgery, assess which LMH-related factors best predict final VA, and document postoperative complications.

## Material and methods

### Study Design and Population

This retrospective interventional study includes medical records of patients with LMH who underwent PPV between 2014 and 2021 at the Centre Hospitalier Universitaire (CHU) de Québec – Université Laval. Approval from the Ethics Committee was obtained, and the study adhered to the tenets of the Declaration of Helsinki. We excluded eyes with macular pseudohole (MPH), Full thickness macular hole (MH), history of ocular trauma leading to LMH formation, wet age-related macular degeneration (AMD), active proliferative diabetic retinopathy (PDR), advanced glaucoma, high myopia (HM) (>4 diopters), history of retinal detachment (RD), and active uveitis. Patients with less than one month of postoperative follow-up and patients with missing preoperative or postoperative best-corrected VA (BCVA) and optical coherence tomography (OCT) data were also excluded.

### LMH SD-OCT Diagnosis

We defined and diagnosed LMH using the OCT diagnostic criteria of Witkin et al. as these were adopted by the International Vitreomacular Traction Study Group.^15^ Anatomic characteristics were collected using SD-OCT (Cirrus HD-OCT; Carl Zeiss Meditec, Inc). Patients diagnosed with LMH were further classified into ERMFs and “true” LMHs according to Hubschman et al.’s consensus.^4^ We analyzed the following parameters using Cirrus’ OCT 512×128 macular cube scan: preoperative central foveal thickness (CFT), average foveal thickness (AFT), minimal foveal thickness (MFT), base diameter and apex diameter of the hole, presence of ERM and ERP, EZ disruption, external limiting membrane (ELM) disruption, presence of intraretinal cysts (IRCs), postoperative MH formation (stages 1, 2, 3 and 4), and foveal profile evolution. As previously described, LMH closure was defined as a reconstituted foveal contour with no retinal splitting.^16^ The preoperative CFT, AFT and MFT were measured automatically by Zeiss’ OCT software. Base and apex diameters were measured manually using Zeiss’ OCT software caliper. Since lamellar macular defects often exhibit asymmetry and irregularity, measurements of base diameter were made to the largest horizontal extent of the intraretinal split, as performed in similar studies.^17,18^

### Surgical procedure

All patients underwent 25-gauge PPV by one of five fellowship-trained vitreoretinal surgeons. ERM peeling with or without internal membrane (ILM) peeling was performed. Phacoemulsification was also combined to the primary PPV in some cases. A conventional peeling technique was performed on all patients, consisting of removing the epiretinal membrane or epiretinal proliferation if present from the surface of the retina using staining agents. Indocyanine green or methylene blue were used as staining agents based on surgeons’ preference. ILM peeling and restaining were done following the removal of ERM or ERP. In exceptional cases where the ILM could not be visualized after extensive ERM peeling, the ILM was not removed. Vitrectomy was completed with a 360° inspection of the peripheral retina using scleral depression, followed by complete fluid-air exchange with or without gas tamponade at the surgeon’s discretion. The gases used for tamponade were either air, sulfar hexafluoride (SF6), or perfluoropropane (C3F8) for larger LMHs, chosen at the discretion of the operating surgeon. Sclerotomies were sutured if they were not self-sealing and leakage occurred. Patients who received gas tamponade (SF6 and C3F8) were instructed to maintain prone positioning following surgically.

### Statistical analysis

We conducted the statistical analysis using SPSS Statistics Version 26 (IBM, Armonk, NY, USA). The normality of continuous variables was tested with Q-Q plots and Shapiro-Wilk tests. Means and standard deviations were used to present normally distributed continuous variables, medians and quartiles [first quartile (Q1), third quartile (Q3)] for non-normally distributed continuous variables, and percentages for categorical variables. We compared preoperative characteristics and outcomes between patients using independent Student’s t-tests for normally distributed continuous variables, Mann-Whitney U tests for non-normally distributed continuous variables, Pearson chi-square tests for categorical variables, and Wilcoxon signed-rank tests for paired comparisons of preoperative and postoperative continuous variables.

To analyze which variables were most predictive of final postoperative BCVA, we built a multiple linear regression model using backwards elimination with an F-to-remove at 0.2. Variables that we considered for inclusion were age, sex, idiopathic LMH, preoperative BCVA, use of phaco- vitrectomy, type of peel (e.g., ERM peel, ILM peel, and/or ERP peel), tamponade agent, staining, LMH closure, preoperative central foveal thickness, lesion type (i.e., ERMF or “true” LMH), LMH base and apex diameters, preoperative OCT characteristics if they were not significantly collinear with lesion type (i.e., ERP, EZ disruption, ELM disruption, IRC, vitreomacular traction (VMT), partial/complete posterior vitreous detachment (PVD), cystoid macular edema (CME), and development of postoperative MH). Unstandardized coefficients with 95% confidence intervals (CI) and standardized coefficients were produced for all variables included in the final model. Baseline demographics including preoperative BCVA, age, and sex were retained in the final model to adjust for these variables. The type of lesion (ERMF vs “true” LMH) and LMH closure were included to assess the impact of these anatomical characteristics on final BCVA. Additionally, we favored the use of phaco-vitrectomy over other lens status variables and preoperative central foveal thickness over other preoperative thickness parameters (i.e., minimal foveal thickness, average foveal thickness) due to their stronger association with final BCVA and to avoid collinearity in the regression model. Statistical significance was set at a p-value of <0.05.

## Results

### Characteristics of the Studied Population

This study included 90 patients. Out of them, 57 (63.3%) were women. The mean age at surgery was 71 ± 8 years. Of these, 51 (56.7%) subjects presented with ERMF, while 39 (43.3%) presented with “true” LMH. Comparisons of baseline characteristics between both types of lesions are presented in **Table 1**. Out of the 39 patients presenting “true” LMH, 28 (71.8%) were subject to an ERP and ILM peeling, 6 (15.4%) had an ERP peeling, and 5 (12.8%) had an ILM peeling. The subset of “true” LMH patients who underwent solely an ILM peeling did not have ERP at presentation. Out of the subjects presenting ERMF, 46 (90.2%) had an ERM and an ILM peeling, while 5 (9.8%) had only an ERM peeling. Following the ERM or ERP peeling, the decision to peel ILM was at the discretion of the surgeon. In the entire cohort, 58 (64.4%) subjects received SF6, 30 (33.3%) received air, and 2 (2.2%) received C3F8. Concomitant phacoemulsification and intraocular lens (IOL) implantation were performed in 24 (26.7%) subjects. The median [Q1, Q3] follow-up period was 14 [6, 29] months.

**Table 1.**
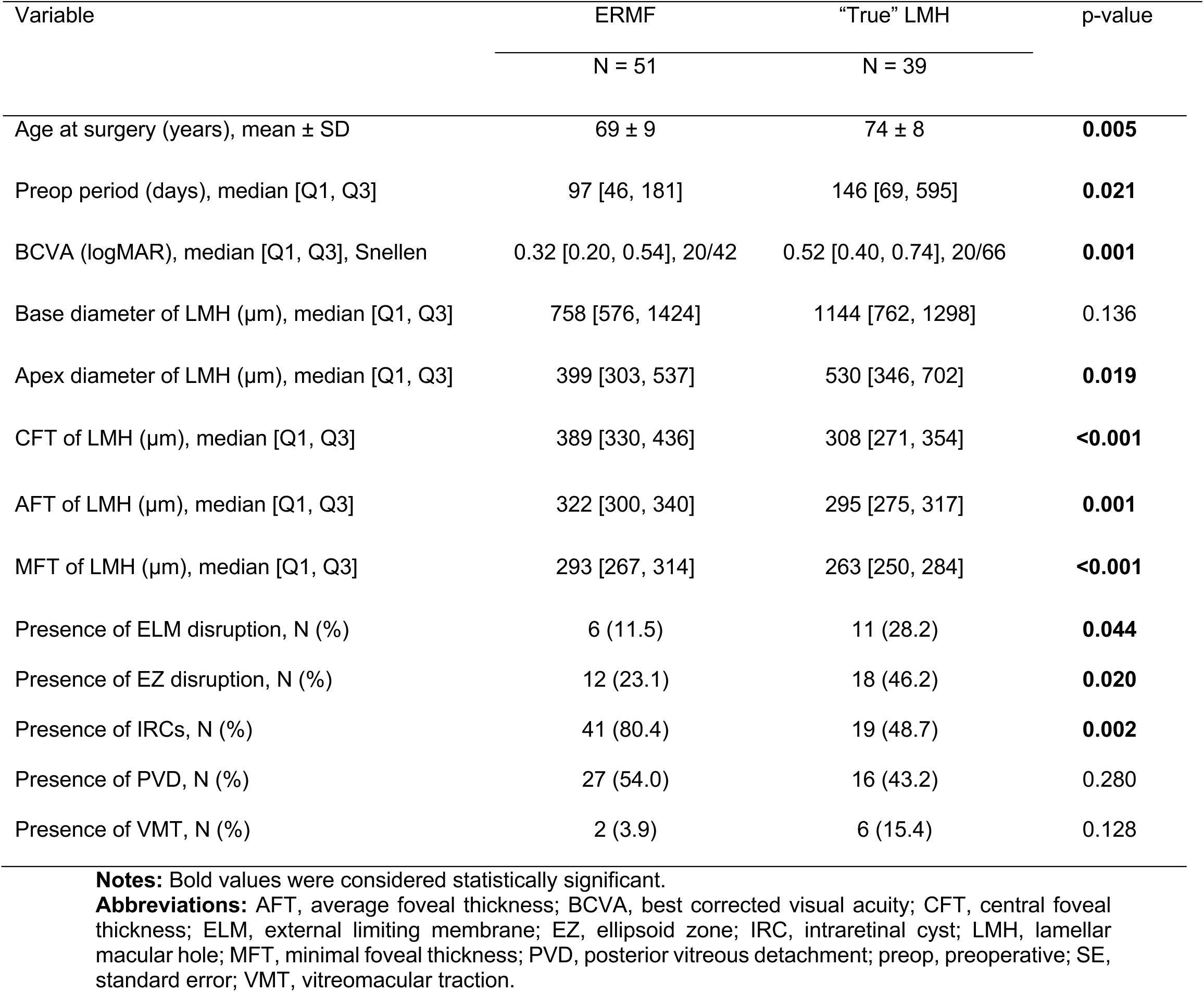
Comparison of Preoperative Clinical and Optical Coherence Tomography Findings of ERMF and “true” LMH Patients.

| Variable | ERMF | “True” LMH | p-value |
| --- | --- | --- | --- |
|  | N = 51 | N = 39 |  |
| Age at surgery (years), mean ± SD | 69 ± 9 | 74 ± 8 | **0.005** |
| Preop period (days), median [Q1, Q3] | 97 [46, 181] | 146 [69, 595] | **0.021** |
| BCVA (logMAR), median [Q1, Q3], Snellen | 0.32 [0.20, 0.54], 20/42 | 0.52 [0.40, 0.74], 20/66 | **0.001** |
| Base diameter of LMH (μm), median [Q1, Q3] | 758 [576, 1424] | 1144 [762, 1298] | 0.136 |
| Apex diameter of LMH (μm), median [Q1, Q3] | 399 [303, 537] | 530 [346, 702] | **0.019** |
| CFT of LMH (μm), median [Q1, Q3] | 389 [330, 436] | 308 [271, 354] | **<0.001** |
| AFT of LMH (μm), median [Q1, Q3] | 322 [300, 340] | 295 [275, 317] | **0.001** |
| MFT of LMH (μm), median [Q1, Q3] | 293 [267, 314] | 263 [250, 284] | **<0.001** |
| Presence of ELM disruption, N (%) | 6 (11.5) | 11 (28.2) | **0.044** |
| Presence of EZ disruption, N (%) | 12 (23.1) | 18 (46.2) | **0.020** |
| Presence of IRCs, N (%) | 41 (80.4) | 19 (48.7) | **0.002** |
| Presence of PVD, N (%) | 27 (54.0) | 16 (43.2) | 0.280 |
| Presence of VMT, N (%) | 2 (3.9) | 6 (15.4) | 0.128 |

**Notes:** Bold values were considered statistically significant.
**Abbreviations:** AFT, average foveal thickness; BCVA, best corrected visual acuity; CFT, central foveal thickness; ELM, external limiting membrane; EZ, ellipsoid zone; IRC, intraretinal cyst; LMH, lamellar macular hole; MFT, minimal foveal thickness; PVD, posterior vitreous detachment; preop, preoperative; SE, standard error; VMT, vitreomacular traction.

### Anatomic Outcomes

Lamellar macular hole closure was achieved in 80 (88.9%) cases. **Figure 1** depicts the preoperative and postoperative OCT images of a patient who achieved complete LMH closure, compared to a patient who did not. The characteristics of patients who did or did not achieve LMH closure are summarized in **Table 2**. In addition to restoring foveal profile, surgery significantly decreased the number of patients presenting IRCs, which went from 60 (66.7%) preoperatively to 32 (35.6%) postoperatively (p<0.001). The number of patients with EZ disruption who achieved restoration of EZ integrity postoperatively was 6 (representing a decrease of 20.7%) (p=0.238), while the number of patients presenting CME decreased from 6 (6.6%) to 5 (5.6%) postoperatively (p=1.000).

**Figure 1.**
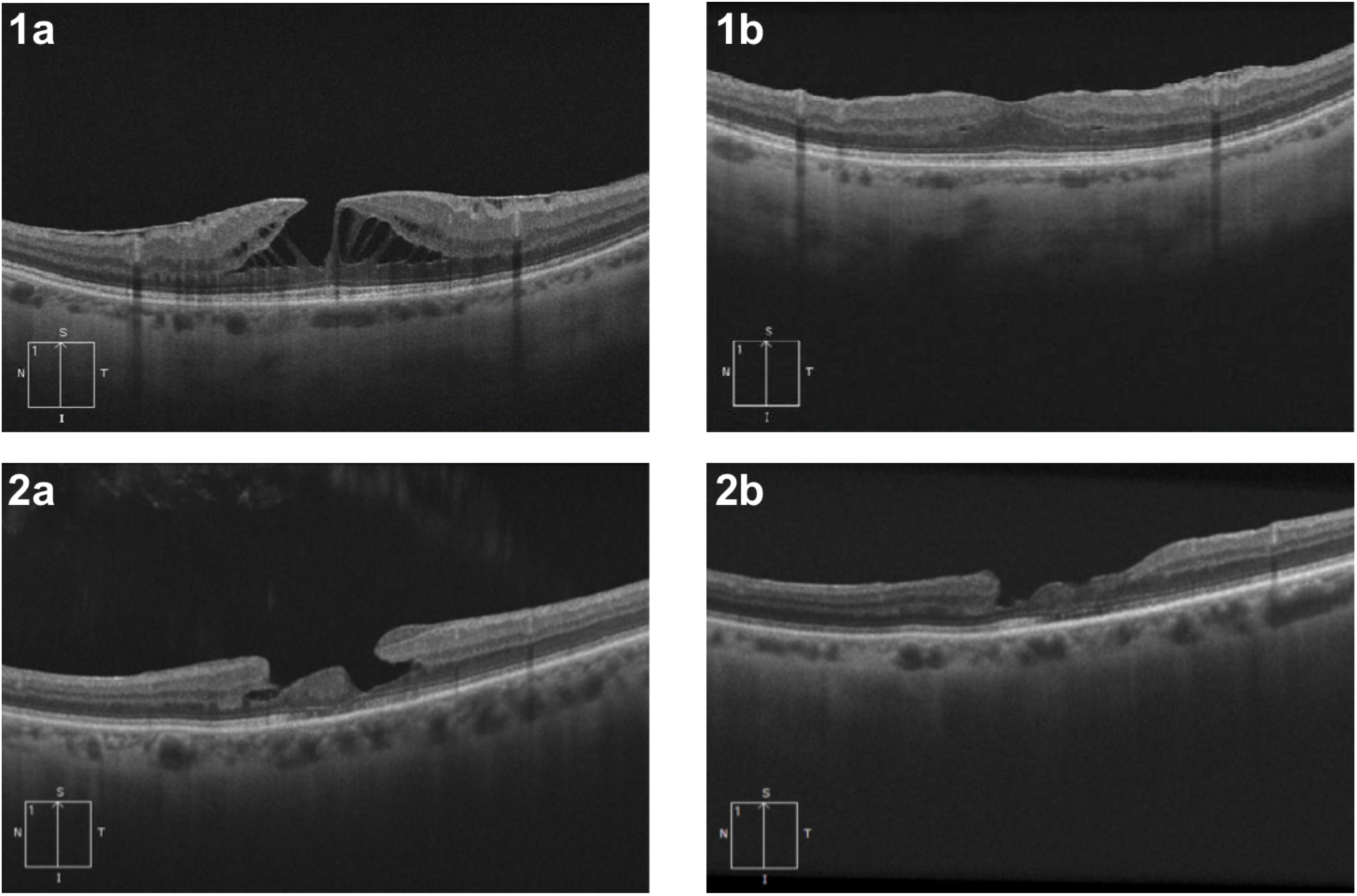
Optical coherence tomography features of cases of lamellar macular hole (LMH) closure and non-closure postoperatively. The patient who achieved LMH closure initially presented with epiretinal membrane foveoschisis (1a). Restauration of the retinal layers was achieved following surgery (1b). The patient who did not achieve LMH closure initially presented with “true” LMH and epiretinal proliferation (2a). Although the epiretinal proliferation was successfully removed during surgery, the patient did not achieve full LMH closure following surgery, as a break in the inner layers of the retina remained.

**Table 2.**
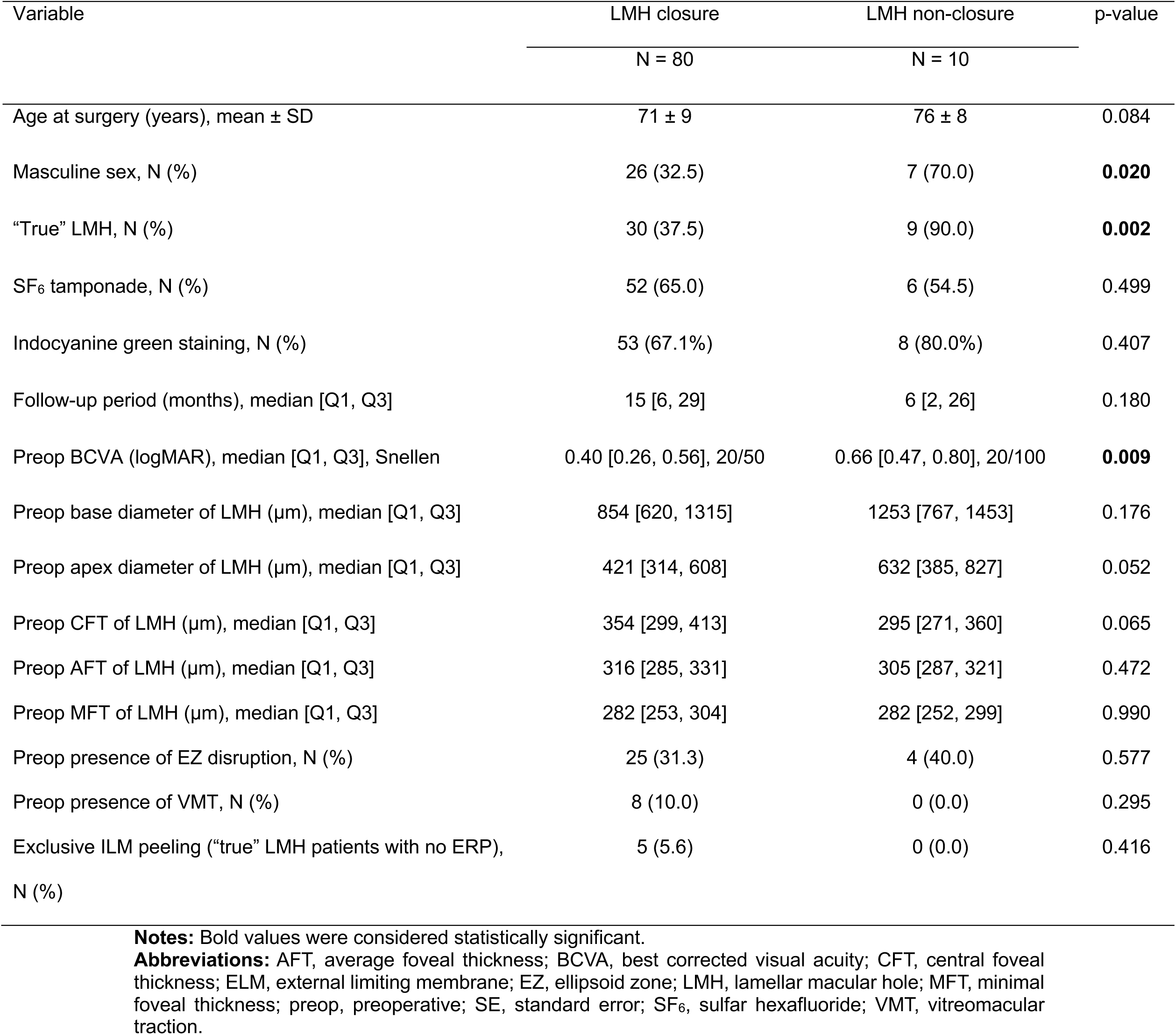
Comparison of patient characteristics between cases which did and did not lead to lamellar macular hole (LMH) closure.

Of the 10 patients who did not achieve LMH closure, 7 (70.0%) of them were men (p=0.020). “True” LMH was associated with a lower rate of LMH closure than ERMF (p=0.002). Subjects who did not achieve LMH closure had significantly worse preoperative BCVA (LMH closure group: 0.40 [0.26, 0.56], Snellen 20/50 vs. LMH non-closure group: 0.66 [0.47, 0.80], Snellen 20/100; p=0.009). No significant association was found between LMH closure and the type of tamponade agents. Patients who achieved LMH closure had higher CFT values, although not reaching statistical significance (LMH closure group: 354 [299, 413], LMH non-closure group: 295 [271, 360], p=0.065). The preoperative apex diameter was smaller in patients who achieved LMH closure (421 [314, 608] μm) than in the ones who did not (632 [385, 827] μm, p=0.052). Age, preoperative base diameter, preoperative presence of ELM disruption, EZ disruption, vitreomacular traction, intraretinal retinal cysts, and PVD were not significantly associated with LMH closure.

When considering the “true” LMH subgroup alone, the proportion of patients with ERP that did not achieve LMH closure (n=9, 100% of LMH non-closure cases) was significantly higher than the proportion of patients with ERP that achieved LMH closure (n=18, 60% of LMH closure cases), highlighting a negative association between the presence of ERP and LMH closure. There was no other preoperative anatomic characteristic associated with LMH closure for “true” LMH cases.

### Functional Outcomes

Surgery significantly improved BCVA from a preoperative median [Q1, Q3] BCVA of (0.42 [0.26, 0.61]) (Snellen: 20/50) to a final postoperative BCVA of (0.31 [0.14, 0.48]) (Snellen: 20/40) (p=0.024). **Table 3** summarizes the factors associated with final BCVA when adjusted for preoperative BCVA in a multiple linear regression model. Variables not included in the model were rejected by the backward elimination method.

**Table 3.** Multiple linear regression model for final best-corrected visual acuity (BCVA) following surgery for lamellar macular hole (LMH) in 90 patients.

| Variable | B | 95% CI | $\beta$ | p-value |
| --- | --- | --- | --- | --- |
| Age at surgery | 0.005 | -0.004, 0.013 | 0.108 | 0.266 |
| Sex | 0.275 | 0.134, 0.415 | 0.358 | **<0.001** |
| Preop BCVA | 0.188 | -0.067, 0.443 | 0.144 | 0.146 |
| Phaco-PPV | -0.281 | -0.437, -0.125 | -0.343 | **<0.001** |
| No ILM peeling | 0.226 | -0.101, 0.552 | 0.129 | 0.173 |
| Exclusive ILM peeling ("true" LMH patients with no ERP) | 0.304 | 0.015, 0.594 | 0.194 | **0.040** |
| SF<sub>6</sub> tamponade | 0.109 | -0.037, 0.255 | 0.141 | 0.142 |
| LMH closure | -0.048 | -0.274, 0.177 | -0.042 | 0.671 |
| Preop CFT of LMH | -0.001 | -0.002, 0.000 | -0.128 | 0.192 |
| Lesion type (ERMF/"true" LMH) | -0.037 | -0.205, 0.131 | -0.050 | 0.665 |
| Preop base diameter of LMH | 0.000 | 0.000, 0.000 | -0.142 | 0.131 |
| Preop presence of EZ disruption | 0.144 | -0.006, 0.295 | 0.182 | 0.060 |
| Preop presence of IRCs | -0.118 | -0.268, 0.032 | -0.150 | 0.123 |

**Notes:** Bold values were considered statistically significant.
**Abbreviations:** AFT, average foveal thickness; BCVA, best corrected visual acuity; B, unstandardized coefficients; $\beta$ , standardized coefficients; CI, confidence interval; CFT, central foveal thickness; ERM, epiretinal membrane; ERMF, epiretinal membrane foveoschisis; ERP, epiretinal proliferation; EZ, ellipsoid zone; ILM, internal limiting membrane; IRC, intraretinal cyst; LMH, lamellar macular hole; PPV, pars plana vitrectomy; preop, preoperative; SF<sub>6</sub>, sulfur hexafluoride.

Multiple linear regression analysis indicated that, placed in order of magnitude of impact, masculine sex (β=0.358, p<0.001) and “true” LMH with no ERP (β=0.194, p=0.040) were significant predictors negatively affecting final postoperative BCVA. On the other hand, phaco-vitrectomy (β=- 0.343, p<0.001) was a significant predictor positively affecting final postoperative BCVA. LMH type, whether “true” LMH or ERMF, was not a significant predictor of postoperative BCVA. All other variables were not significantly associated with final postoperative BCVA.

### Postoperative Complications

Preoperatively, 61 (67.0%) patients were phakic, among whom 24 (39%) underwent phaco- vitrectomy. The number of subjects who presented cataracts increased significantly after surgery (p<0.001), rising from 53 (58.9%) to 80 (88.9%). After surgery, one patient (1.1%) developed uveitis. MH occurred in 5 (5.5%) cases postoperatively. **Figure 2** depicts preoperative and postoperative OCT images of a patient who experienced an MH postoperatively and subsequently underwent a secondary vitrectomy. **Table 4** summarizes the characteristics of the patients who experienced postoperative MH development.

**Figure 2.**
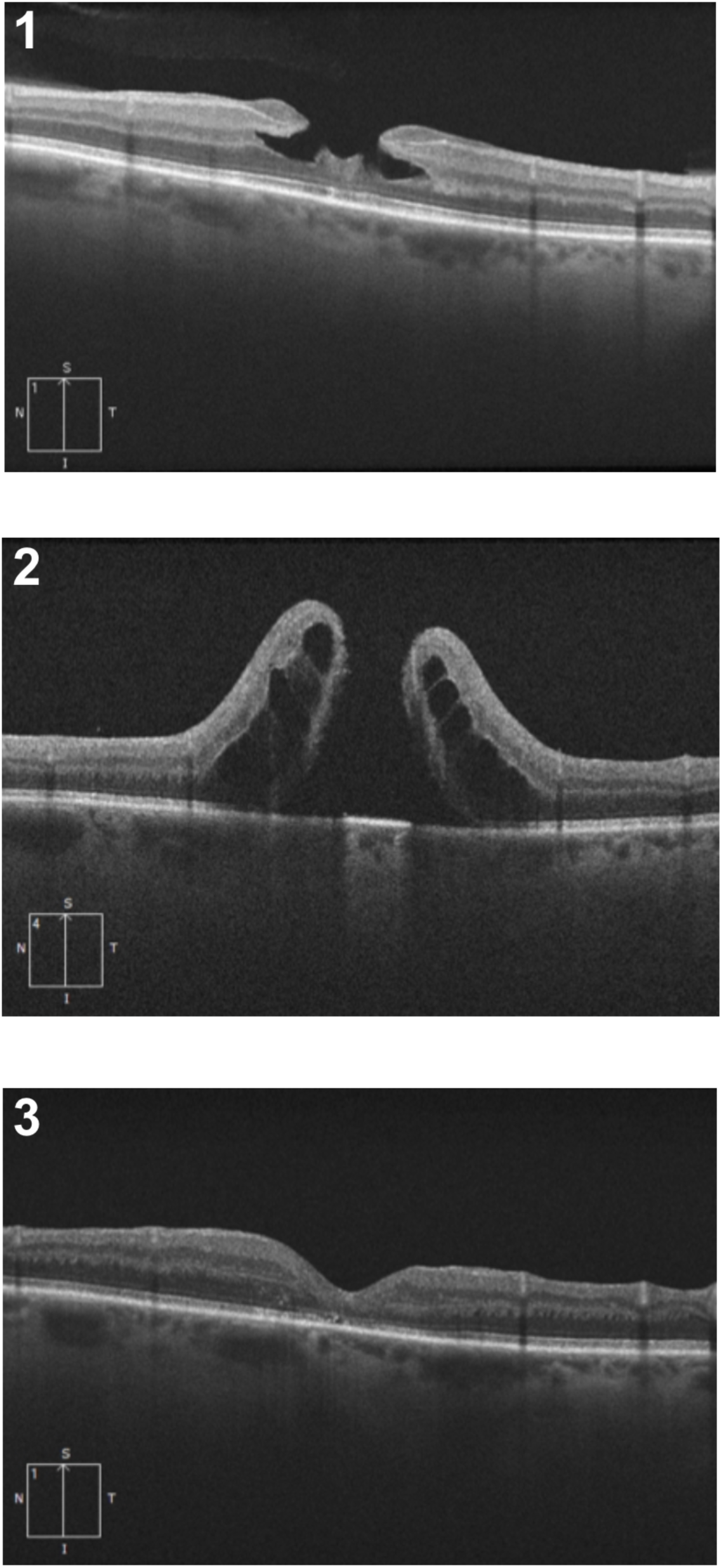
Optical coherence tomography features of a “true” lamellar macular hole patient undergoing surgery and experiencing postoperative full-thickness macular hole postoperatively. The patient initially presented with idiopathic “true” lamellar macular hole and epiretinal proliferation of medium reflectivity (1). Posterior vitreous detachment had to be induced surgically. The patient received sulfar hexafluoride (SF6) tamponade and was instructed to maintain prone positioning for three days following surgery. The patient underwent a secondary revision surgery 36 days postoperatively as he developed stage 4 MH that was discovered during his two-week postoperative appointment (2). Following the second surgery, normalisation of the foveolar profile was acheived in the patient (3).

**Table 4.** Clinical data and optical coherence tomography (OCT) characteristics of postoperative macular hole cases.

| Patient/<br>Sex | LMH type | BCVA (logMAR, Snellen) |  | Surgical modalities |  |  | Preop OCT characteristics |  |  |
| --- | --- | --- | --- | --- | --- | --- | --- | --- | --- |
| | | Preop | Postop (final) | Peeling | Tamponade | Prone<br>positionning | CFT/AFT<br>($\mu\text{m}$) | Base/apex<br>diameter<br>($\mu\text{m}$) | Preop EZ<br>disruption |
| 1/M | “true” LMH | 1.3, 20/400 | 0.64, 20/80<sup>-2</sup> | ERP + ILM | SF<sub>6</sub> | No | 289/305 | 1246/233 | Yes |
| 2/M | “true” LMH | 0.92, 20/150<sup>-2</sup> | 0.88, 20/150 | ERP + ILM | SF<sub>6</sub> | No | 354/305 | 1514/347 | Yes |
| 3/F | “true” LMH | 0.28, 20/40<sup>+1</sup> | 0.28, 20/40<sup>+1</sup> | ERP + ILM | SF<sub>6</sub> | Yes | 271/313 | 1303/780 | Yes |
| 4/M | “true” LMH | 0.76, 20/100<sup>-3</sup> | 0.66, 20/30<sup>+2</sup> | ERP + ILM | Air | Yes | 293/309 | 1432/633 | No |
| 5/M | “true” LMH | 0.62, 20/80<sup>-1</sup> | 1.28, 20/400<sup>+1</sup> | ERP + ILM | SF<sub>6</sub> | Yes | 379/528 | 1200/899 | No |

**Abbreviations:** AFT, average foveal thickness; AMD, age-related macular degeneration; BCVA, best corrected visual acuity; CFT, central foveal thickness; ERP, epiretinal proliferation; EZ, ellipsoid zone; GA: geographic atrophy; ILM, internal limiting membrane; LMH, lamellar macular hole; OCT, optical coherence tomography; postop, postoperative; preop, preoperative; SF<sub>6</sub>, sulfur hexafluoride.

Postoperative MH was associated with “true” LMH as all 5 patients who developed the complication were “true” LMH subjects (p=0.008). Sex was also associated with postoperative MH as 4 (80.0%) subjects who developed MH were males (p=0.036). Patients with postoperative MH had a significantly worse preoperative median [Q1, Q3] BCVA (postoperative MH group: 0.76 [0.45, 1.11], no postoperative MH group: 0.42 [0.26, 0.56], p=0.033). Other parameters were not significantly associated with postoperative MH.

## Discussion

The purpose of this study was to analyze the functional and anatomic outcomes of LMH surgery using the latest classification.^4^ Results indicate that LMH surgery is an effective treatment for LMH, resulting in high rates of closure (88.9%) and improved visual outcomes (median gain of 1 line on the Snellen chart), which confirms previous meta-analysis findings.^19^ Whether ERMF and “true” LMH have different surgical outcomes was unclear. Previous studies showed heterogenous results; with some indicating a postoperative discrepancy between the two entities and some not reporting differences.^12, 19–24^ Hubschman et al.’s reclassification implied that LMH and ERMF might have different surgical outcomes as they may follow distinct pathological pathways.^4^ Thus, the present study also aimed at investigating that matter. LMH surgery had positive functional outcomes for both ERMF and “true” LMH as lesion type was not significantly associated with postoperative final BCVA when adjusted for confounding variables. However, “true” LMH cases represented 90.0% of the cases which did not achieve LMH closure and 100% of the cases which developed postoperative MH.

Venkatesh et al. associated the presence of ERP in LMH with lower visual acuity, larger LMH size, thinner residual retinal tissue, larger EZ disruption, and larger inner segment/outer segment (IS/OS) defects.^23^ The results of the present study align with the latter, as cases presenting with ERP had worse baseline BCVA, a higher incidence of ELM and EZ defects, a thinner central fovea, a higher LMH non-closure rate, as well as a higher incidence of postoperative MH. “True” LMH patients were older and waited longer to undergo surgery, which may suggest that “true” LMH could be a subsequent manifestation or a more advanced stage of the condition, as proposed by Lee et al.^7^ Omoto et al. reported that the presence of ERP was not significantly related to postoperative VA, which contrasts with the findings of the current study.^20^ When adjusted for preoperative BCVA, “true” LMH patients without ERP had worse final BCVA in the present report. These “true” LMH cases had solely undergone an ILM peeling.

Patients with higher CFT values had better anatomic results, as they presented a milder loss of foveal tissue. While not reaching statistical significance, EZ disruption was related to worse final postoperative BCVA, and subjects who did not achieve LMH closure tended to have larger base diameter and apex diameter of the LMH. These findings emphasize the importance of evaluating anatomic foveal features before proceeding with surgery.

Moreover, a significant worsening predictor of both functional and anatomic outcomes was masculine sex. Age at presentation was not significantly different between sexes, neither was the frequency of a specific lesion type.

Haave et al. suggested that if cataract is present, combining phaco-vitrectomy during surgical intervention could optimize functional outcomes.^25^ Coassin et al. conducted a retrospective study of patients who underwent surgical treatment for symptomatic LMHs where pseudophakic patients exhibited better outcomes than phakic patients.^16^ The current study’s results are consistent with those findings as postoperative BCVA was significantly improved with phaco-vitrectomy. Haave et al. further reported that gas tamponade should be avoided as patients who were exclusively administered a balanced salt solution (BSS) achieved the most favorable outcomes.^25^ However, no significant relationship was found between air, SF6 or C3F8 tamponade and postoperative outcomes in the present study. These results suggest that similarly to what was recently reported for idiopathic macular holes in Dervenis et al.’s systematic review and meta-analysis, the choice of tamponade does not affect visual outcomes or closure rates in lamellar macular hole surgery.^26^

All postoperative FTMH cases were “true” LMH cases. Other studies have also reported the occurrence of postoperative FTMH in “true” LMH cases, as the ERP often present in such cases is more challenging to peel.^27, 28^ Thus, the fovea sparing and the flap embedding peeling techniques have emerged as alternatives for treating LMH.^29,30^ The rationale behind them is to avoid peeling the edges of the LMH which are oftentimes connected to the ERP. Such studies have reported positive outcomes and no postoperative FTMH, but further comparatives studies are warrented to establish the superiority of a peeling technique over the other. ^29,30^

Limitations of this study include its retrospective nature. The surgery was performed by multiple surgeons, which might induce heterogeneity in outcomes. However, surgical technique did not differ between surgeons. The study’s strengths include its relatively large cohort size (n=90), making it one of the largest in comparison to similar research, and the largest using the latest classification of LMH.^4^ The study also examined how the OCT subtype of LMH and the used surgical technique affected visual and anatomical outcomes, which was not done in previous studies. Furthermore, the study benefits from a relatively long median follow-up period, enabling comprehensive evaluation of surgical outcomes over an extended duration. Finally, various controversial factors that could influence surgical outcomes were examined, providing valuable insights of the key considerations for evaluating the likelihood of surgical success.

## Conclusion

In conclusion, this study sheds light on the characteristics and outcomes of patients undergoing surgery for “true” LMH and ERMF. It further supports the effectiveness of primary vitrectomy as a viable treatment option for LMH patients, particularly those undergoing concomitant cataract treatment. “True” LMH and ERMF patients had significantly different anatomic characteristics at presentation. Although functional outcomes did not significantly differ between the two groups, anatomic outcomes were worse in “true” LMH. Therefore, specific considerations should be given to OCT biomarkers and patients presenting “true” LMH to optimize surgical outcomes and limit postoperative complications.

## Data Availability

All data produced in the present study are available upon reasonable request to the authors.

## Acknowledgments

The patients included in this study for enabling advancement in the field.

